# COVID-19 disease and economic burden in adults in six Latin American Countries before nationwide vaccination program: Ministry of Health databases assessment and systematic literature review

**DOI:** 10.1101/2024.10.31.24316541

**Authors:** Natalia Espinola, Cecilia I. Loudet, Rosario Luxardo, Carolina Moreno, Moe H. Kyaw, Julia Spinardi, Carlos Fernando Mendoza, Carolina M. Carballo, Ana Carolina Dantas, Maria Gabriela Abalos, Jamile Ballivian, Emiliano Navarro, Ariel Bardach

**Affiliations:** Department of Health Technology Assessment and Health Economics, Institute for Clinical Effectiveness and Health Policy (IECS), Ciudad de Buenos Aires, Argentina; Centro de Investigaciones Epidemiológicas y Salud Pública (CIESP-IECS). CONICET, Ciudad de Buenos Aires, Argentina; Vaccine Scientific Affairs, Pfizer Inc., New York, NY, United States; Vaccine Medical Affairs, Emerging Markets, Pfizer Inc., Itapevi, Brazil; Vaccine HTA, Value and Evidence, Pfizer Inc., Mexico City, Mexico; Vaccine Medical Manager, Pfizer Inc., Buenos Aires City, Argentina

**Author notes:** Corresponding author: Natalia Espinola, BSc., MSc., Health Technology Assessment and Health Economics Department, Institute for Clinical Effectiveness and Health Policy (IECS), Doctor Emilio Ravignani 2024, Buenos Aires - Argentina.

**Keywords:** Adults, COVID-19, Disease Burden, Economic Burden, Latin American

## Abstract

**Introduction and objective:** The COVID-19 pandemic had a significant disease burden on the healthcare system worldwide. There is limited reliable national data on COVID-19 associated morbidity, mortality, and healthcare costs in Latin America. This study explored the COVID-19 disease and economic burden in Argentina, Brazil, Chile, Colombia, Mexico, and Peru in the pre-vaccination period.

**Methods:** We analyzed national databases and conducted a systematic review of the published literature on COVID-19 in adults aged 18 years and above. We reported cases, death rates, years of life lost, and excess mortality associated with COVID-19 in pre-vaccination period. In addition, we used a cost of illness analysis to estimate the direct medical costs associated with COVID-19. All costs are reported in US dollars as of 2023.

**Results:** Before the national COVID vaccination program rollout, the average incidence rate of COVID-19 in adults aged 18 and above was 6,741 per 100,000 persons. Approximately 91% of the cases were mild, 7% were moderate/severe, and 2% were critical. Among 2,201,816 hospitalized cases, 27.8% were in intensive care, and 17.5% required mechanical ventilation. The country-specific data varied greatly across Latin American countries, with hospitalization admission: 469-18,096 per 100,000, excess mortality: 76-557 per 100,000, and years of life lost: 241,089-3,312,346. Direct medical costs ranged from US$258 million to US$10,437 million, representing approximately 2%-5% of national health expenditure.

**Conclusion:** COVID-19 imposed a substantial public health and economic burden on Latin American countries during the pre-vaccination period. The findings should help policymakers to make informed decisions and allocate resources effectively to improve national strategies around surveillance, preventive and treatment strategies to control the spread of COVID-19 disease in the future.

## Introduction

The COVID-19 pandemic has placed substantial strain on the healthcare systems worldwide. Latin America has been particularly hard-hit, with several countries reporting high cases and death tolls. The region has accounted for approximately 15% of global cases and 28% of global deaths [1]. Beyond the known health risks to the population, the COVID-19 pandemic has had significant economic repercussions for the health system and society in a region already facing structural difficulties. This impact on the region has been significant, particularly for vulnerable groups such as low-income families, racial minorities, and older adults, those with high job loss and business closures, and among young parents and those with lower incomes [2].

The roll-out of COVID-19 vaccination has been a significant public health effort worldwide. Governments, healthcare organizations, and pharmaceutical companies have collaborated to develop, manufacture, and distribute vaccines at an unprecedented pace. The roll-out of COVID-19 vaccines has been a complex process involving prioritizing high-risk populations, ensuring equitable access to vaccines, and addressing vaccine hesitancy. Despite these challenges, many countries have made significant progress in vaccinating their populations. The success of the vaccine roll-out was crucial for controlling the spread of COVID-19. How to sustain the vaccination program, considering its impact on the community in the post-pandemic scenario, is still a gap to be understood. In this context, the main purpose of this study was to estimate the disease burden of COVID-19 in the adult population before vaccination and the costs for the healthcare system in six countries of Latin America region to inform public health policies.

## Materials and methods

To estimate the burden of disease and perform cost analyses in adults from six Latin American countries (Argentina, Brazil, Chile, Colombia, Mexico, and Peru), we performed a comprehensive review of official statistics databases (such as surveillance systems, national records of the cause of death, and hospitalization when available). We considered the period before COVID-19 vaccination (from the inception until 1st June 2021-Argentina, Brazil, Colombia, Mexico, Peru-; 1st February-Chile) and that were publicly available. This timeframe was determined considering that there was no nationwide vaccination program in place, meaning that no more than 30% of the population had received a single dose of the vaccine and the first two waves of the pandemic were included. When required, we contacted the Ministries of Health officers to access COVID-19 databases to estimate the outcomes of interest for this study. The World Health Organization database was also used for this analysis. Supplementary Material 1 provides information on the sources, criteria related to the official databases, and flowcharts used to determine the population included in the final analysis for each country. We supplemented the data through a systematic literature review when official databases needed more information.

### Inclusion and exclusion criteria

This analysis only included studies that provided data from the adult general population (≥18 years) diagnosed with COVID-19 in Argentina, Brazil, Chile, Colombia, Mexico, and Peru in the pre-vaccination period. We searched the data and stratified the population by sex (female-male) and age (18–49, 50–64, and ≥ 65 years). Also, we searched data on individuals with certain comorbidities (hypertension, diabetes, obesity, chronic kidney disease, asthma, chronic obstructive pulmonary disease, cardiovascular disease, and other forms of immunosuppression). We included surveillance studies, prevalence cohorts, ministerial reports, WHO reports, budget impact analyses, and economic or direct cost evaluations using a microeconomic approach. Randomized clinical trials were excluded from the study. We also excluded reports from Chile after February 1, 2021, and reports from Argentina, Brazil, Colombia, Mexico, and Peru after June 1, 2021, to comply with the established pre-vaccination period.

### Design of studies included

We included studies with any epidemiological or economic design (whose full text was Spanish, English, or Portuguese). Epidemiologically relevant surveillance reports for the selected calendar period were also assessed. We also included observational studies such as cohort, case-control, case series, economic evaluations, and cost or budget impact studies.

Study selection was performed using COVIDENCE, a web-based platform designed for the systematic review process. The authors of the articles were contacted when necessary to obtain missing or supplementary information. Previously piloted in five studies, a predesigned general data extraction form was used. Disagreements were resolved by consensus of the entire team.

### Risk of bias

The risk of bias in observational studies was assessed using a checklist developed by the US National Heart, Lung, and Blood Institute, which classifies studies as having a high risk of bias (poor), uncertain (fair), and low risk of bias (good). For the assessment of cohort and cross-sectional studies, the tool comprises 14 items, while nine items apply to case series studies. The assessments were independently performed by peer reviewers from the research team. Discrepancies were resolved by a consensus of the entire team. Several potential limitations of the literature review include incomplete data in published studies and MoH reports, heterogeneity and quality of available data, and publication bias due to the inclusion of only one geographical region in the review. Supplementary Material 2 shows the literature search and flowchart of the included studies.

The primary health outcomes analyzed were cases defined by the National Health Ministries, deaths, and types of hospitalizations by severity stratified by age group and sex. According to WHO Clinical Practice Guides for the Management of COVID-19 published in 2021, COVID-19 was classified into different categories: mild, moderate, severe, and critical.[3] Firstly, mild COVID-19 is identified in cases requiring symptomatic treatment and not presenting viral pneumonia or hypoxia. On the other hand, cases with non-severe pneumonia are considered moderate COVID-19 cases, and the designation of severe COVID-19 is reserved for patients with pneumonia of considerable severity. Finally, COVID-19 is defined as critical to those patients who present with Acute Respiratory Difficulty Syndrome (ARDS). However, in the analysis of the databases, it was found that the countries did not use this classification for COVID-19 cases, but rather classified them according to the need and type of hospitalization. Therefore, to homogenize the information and use the WHO categorization, the aforementioned definitions were used. However, the definitions of moderate and severe are diffuse and very difficult to categorize according to the type of hospitalization, so we had to unify these two categories into one called moderate and severe.

We also estimated the years of life lost (YLLs), potential years of life lost, disability-adjusted life years (DALYs), and excess mortality (EM). YLLs are defined as the difference between an individual’s age at death and their life expectancy, indicating how many years that person could have lived if they had not died prematurely [4]. To identify these outcomes, we conducted a systematic review and prioritized studies with the widest analysis period (aligned with our study’s timeframe), and reported both total and sex-disaggregated YLLs. We also reported the overall YLLs and age-standardized YLLs (further details are available in Supplementary Material 3). The WHO database also estimated the excess mortality attributed to COVID-19 (Supplementary Material 4). The WHO defines excess mortality due to COVID as the difference between the number of observed deaths from all causes during a given period, and the number of expected deaths from the same period, based on historical data from recent years [5].

The economic burden per patient associated with COVID-19 was calculated using a cost of illness analysis through the micro-costing approach [6]. The perspective of the analysis was the healthcare system. This method consisted of identifying health resources, rates of use, and quantities, then assigning unit costs to each resource.

For the identification of healthcare resources and their use rates, we relied on the WHO guide on managing COVID-19 [7], supplemented with the opinion of local experts with extensive experience in managing COVID-19 patients in ICU settings, which can provide valuable insights into rates of use in real life. Only direct medical costs were considered in this analysis, including drug acquisition and administration costs, disease monitoring (laboratory tests and imaging), hospitalization (General Ward, ICU, and ICU with mechanical ventilation), and medical consultation. We assumed that the rates of use and quantities of most healthcare resources (consultations, laboratory examinations, diagnostic tests, and medications) do not differ between countries, since they are based on good clinical practice, except in the case of length of hospital stay, where we differentiate between countries using information obtained from literature review and indirect estimates through the benefit transfer method. [8][9][10] With this information it was possible to estimate the duration of hospitalization in general wards, ICU without mechanical ventilation, and ICU with mechanical ventilation, according to the level of severity. Supplementary Material 5 details the length of stay in days and the rate of use of healthcare resources based on case severity.

Unit healthcare cost databases were used for each country,[11–14] except for Mexico and Peru, where costs had to be estimated indirectly due to the lack of country-specific information. Drug costs were collected from different sources, depending on the country [15–17]. In the case of Mexico, the drug costs were calculated by indirect estimation using the benefit transfer method.

[8] Details of the unit cost approach and information by severity and country are shown in Supplementary Material 6.

The results are presented as the cost per patient associated with COVID-19, according to the severity of the disease and the total cost faced by the country (multiplying the cost per patient by the number of cases in the country). The costs were updated for inflation using the Consumer’s Price Index obtained from the official pages of each country to June 2023 [18–22]. Costs are reported, based on each country’s nominal exchange rate, in US dollars for 2023 [23–26].

Argentina: ARS/USD $350.00/1, Brazil: BRL/USD $4.90/1, Chile: CLP/USD $855.66/1, Colombia: COP/USD $4,066.89/1, Mexico: MXN/USD $16.98/1, Perú: PEN/USD $3.69/1.

In addition, we explored the disease and economic burden of other preventable diseases (Influenza and Invasive Pneumococcal) before vaccine introduction in the National Immunization Program (NIP) for reference. It is essential to highlight that disease comparisons are inadequate due to differences in burden, epidemiological settings, age prevalence, surveillance system, and agent characteristics, among others. We retrieved data from 2001 to 2017 from ministerial reports from every country and the Surveillance Network System of Agents Responsible for Bacterial Pneumonia and Meningitis (SIREVA). This period was selected based on the vaccination timelines for each country. A complementary literature search was conducted using the PubMed and Lilacs databases to estimate the costs associated with the selected diseases (See Supplementary Material 7). Studies of the economic burden or direct medical costs were identified for each vaccine-preventable disease in Argentina, Brazil, Chile, Colombia, and Peru.

Regarding other vaccine-preventable diseases such as Influenza and *S. pneumoniae* pneumonia, we also assessed health outcomes in adults aged 18 years and above before introducing vaccines for these conditions in the national immunization schedule. The search methodology is described in Supplementary Material 8.

### Statistical Analysis

The overall incidence rates for cases, deaths, and hospitalizations (per 100,000 persons) were extracted from the Ministry of Health (MoH) database for each country (see Table S1 in Supplementary Material). Regarding YLL, age-standardized estimates for people over 20 years of age were derived from data from the Ministry of Health (MoH) database and information extracted from the included studies in the systematic review (Supplementary Material 3). Excess mortality per 100,000 persons, encompassing all age groups in the study population, was also assessed using data from the studies included in the systematic review. The population information for estimating excess mortality rates was sourced from the World Health Organization (WHO) database. Descriptive statistics, including frequency and percentage, were employed. A subgroup analysis was performed based on age (18-49, 50-64, and ≥65 years) and sex. Data were analyzed with R software (4.2.1). Table S18 Supplemental Material 9 shows the data population by country.

## Results

The number of COVID-19 cases diagnosed by the health system ranged between 700,000 (Chile) and 11,000,000 (Brazil). The highest incidence rate of COVID cases was observed in patients under 65 years of age in all countries (Table 1). According to the WHO definition of severity, approximately 90% of the cases were mild, 7% were moderate and severe, and 3% were critical. Generally, males had a higher incidence rate than females in the selected countries (nearly 7 percentage points difference), except for Colombia. Figure 1 presents the daily confirmed COVID-19 cases per 100,000 individuals in the six countries, encompassing both waves. Chile captured the early second wave, while Argentina highlighted its peak.

**Table 1.** COVID-19 cases according to severity and incidence rates per 100,000 persons by country, sex, and age.*

| Indicators | Argentina | Brazil | Chile | Colombia | Mexico | Peru |
| --- | --- | --- | --- | --- | --- | --- |
| COVID-19 All cases, N (%) | 3,711,250 (100) | 11,956,157 (100) | 728,812 (100) | 3,240,355 (100) | 2,363,214 (100) | 1,867,034 (100) |
| Mild n (%) | 3,544,077 (95) | 10,459,717 (88) | 675,039*(93) | 3,133,423* (97) | 1,898,410 (80) | 1,768,939 (94) |
| Moderate and Severe n (%) | 133,339 (4) | 836,398 (7) | 35,236* (5) | 69,900* (2) | 427,694 (18) | 86,796 (5) |
| Critical n (%) | 33,834 (1) | 478,171 (4) | 18,537 (2) | 33,503* (1) | 37,110 (2) | 11,299 (1) |
| Missing data | 0 | 181,871 (1) | 0 | 0 | 0 | 0 |
| Incidence (both sex) | 11,490 | 7,507 | 4,865 | 8,684 | 2,700 | 8,225 |
| 18-49 yrs | 12,362 | 9,635 | 5,122 | 8,712 | 2,486 | 7,719 |
| 50-64 yrs | 11,648 | 3,559 | 4,969 | 8,680 | 3,248 | 9,789 |
| >=65 yrs | 7,886 | 2,968 | 3,724 | 8,527 | 3,039 | 8,696 |
| Incidence Female | 11,210 | 7,154 | 4,767 | 8,852 | 2,597 | 7,821 |
| 18-49 yrs | 12,520 | 9,483 | 5,102 | 9,029 | 2,466 | 7,471 |
| 50-64 yrs | 11,318 | 3,220 | 4,802 | 8,814 | 3,044 | 9,154 |
| >=65 yrs | 6,815 | 2,518 | 3,564 | 7,985 | 2,589 | 7,754 |
| Incidence Male | 11,594 | 7,874 | 4,965 | 8507 | 2,811 | 8,647 |
| 18-49 yrs | 12,035 | 9,779 | 5,143 | 8,391 | 2,506 | 7,974 |
| 50-64 yrs | 11,849 | 3,933 | 4,973 | 8,534 | 3,478 | 10,445 |
| >=65 yrs | 9,092 | 3,569 | 3,922 | 9,204 | 3,575 | 9,781 |

Note: Incidence rate Missing data: Argentina (0.8- 1.7%) Brazil ( 0.01- 0.1% ), Chile, Colombia, Mexico and Peru (0%). Severity based on the definition of disease severity by WHO on the Living Guidance for clinical management of COVID-19 (WHO). Missing data: Brazil (1.5%), Argentina, Chile, Colombia, Mexico and Peru (0%). \* Values estimated from published studies.(Power BI Report )
\*From February 2020 until 1st February 2021 for Chile (12 months), 1st June 2021 for Argentina, Brazil, Colombia, Mexico, and Peru (16 months)

**Figure 1.**
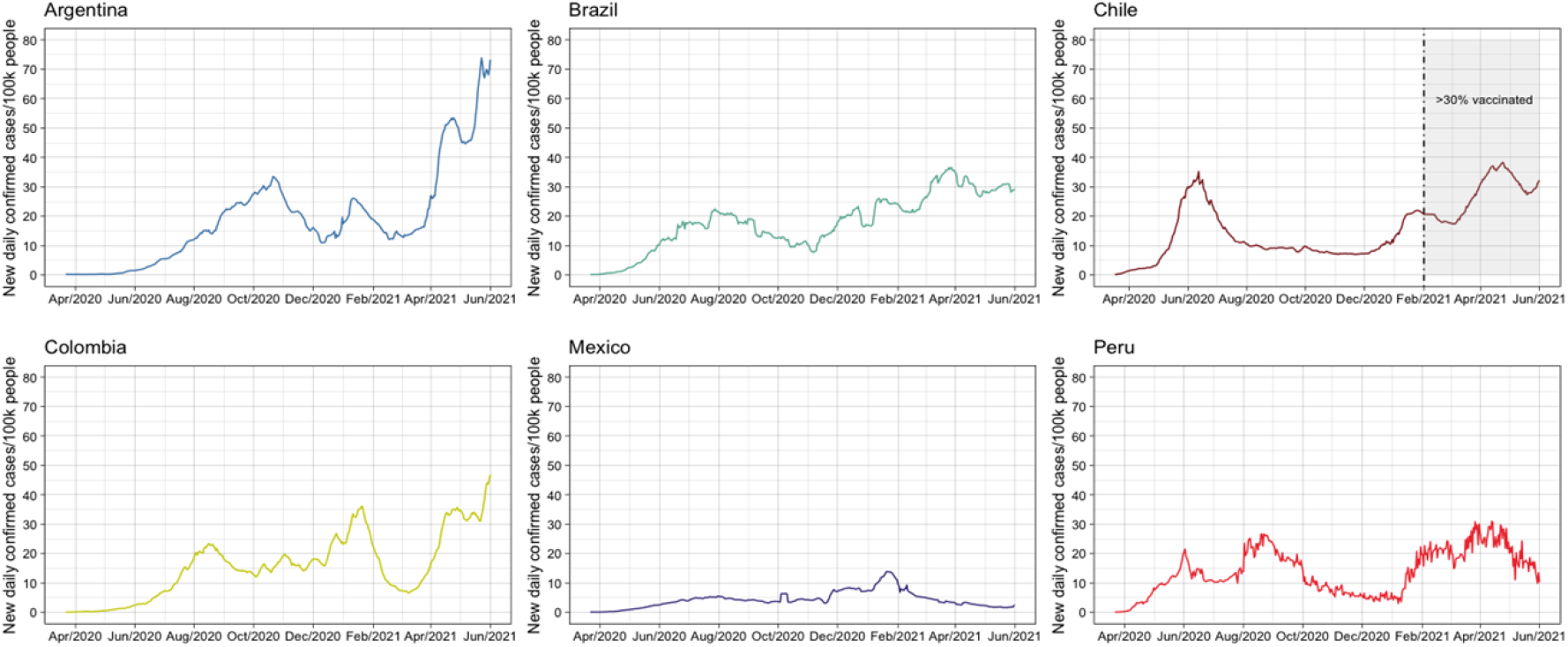
Daily new confirmed COVID-19 cases per 100.000 persons. Notes: elaborated from open-source data from Our World in Data.[27] and normalized to 100.000 persons by country.

### Hospitalizations

Table 2 summarizes the hospitalized COVID-19 cases (sex and age), critical care admission, and hospitalized cases with mechanical ventilation by country. Overall, 9.2% of cases required hospitalization, with a heterogeneous distribution across countries. The majority of hospitalized cases were male (56%) and over 65 years old (39%). A similar distribution was observed in the critical care setting in all countries except Mexico and Peru, where the largest proportion was represented by the age group of 50–64 years. Sex- and age-stratified patients under critical care are summarized in Table S19A in the Supplementary Material. Less than 20% of the patients required mechanical ventilation. Sex and age-stratified patients under mechanical ventilation are summarized in Table S19B of Supplementary Material.

**Table 2.**
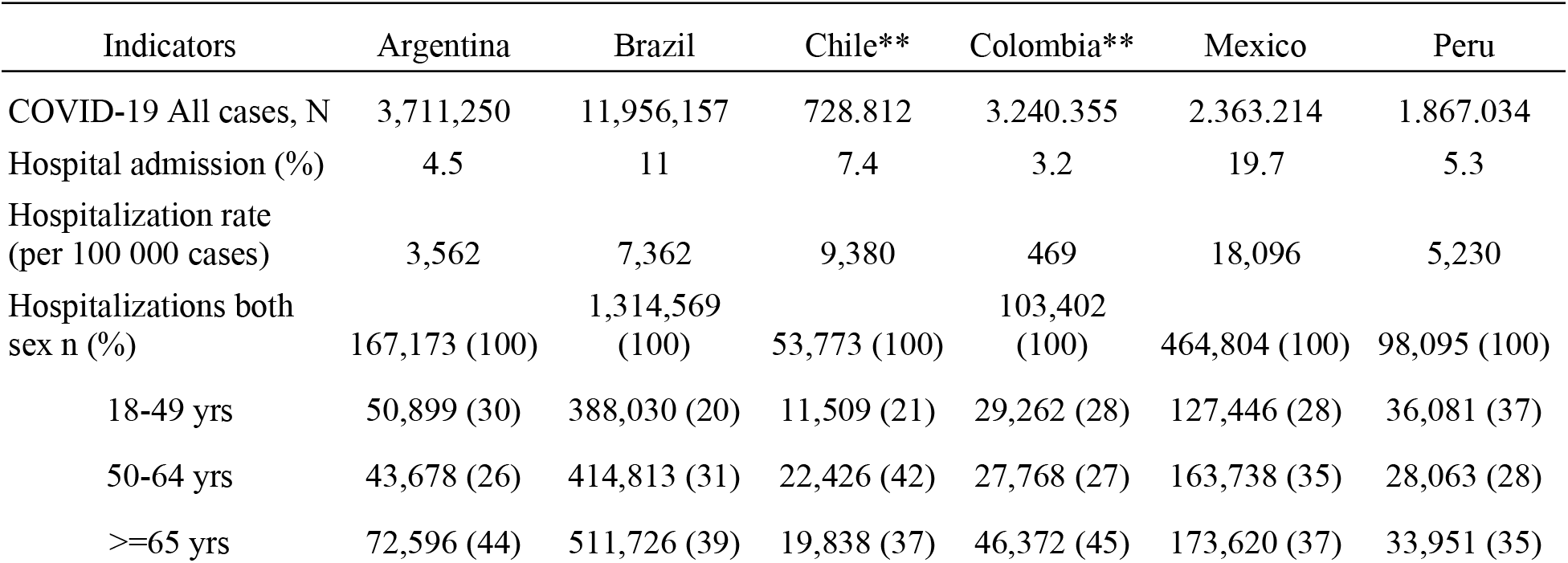
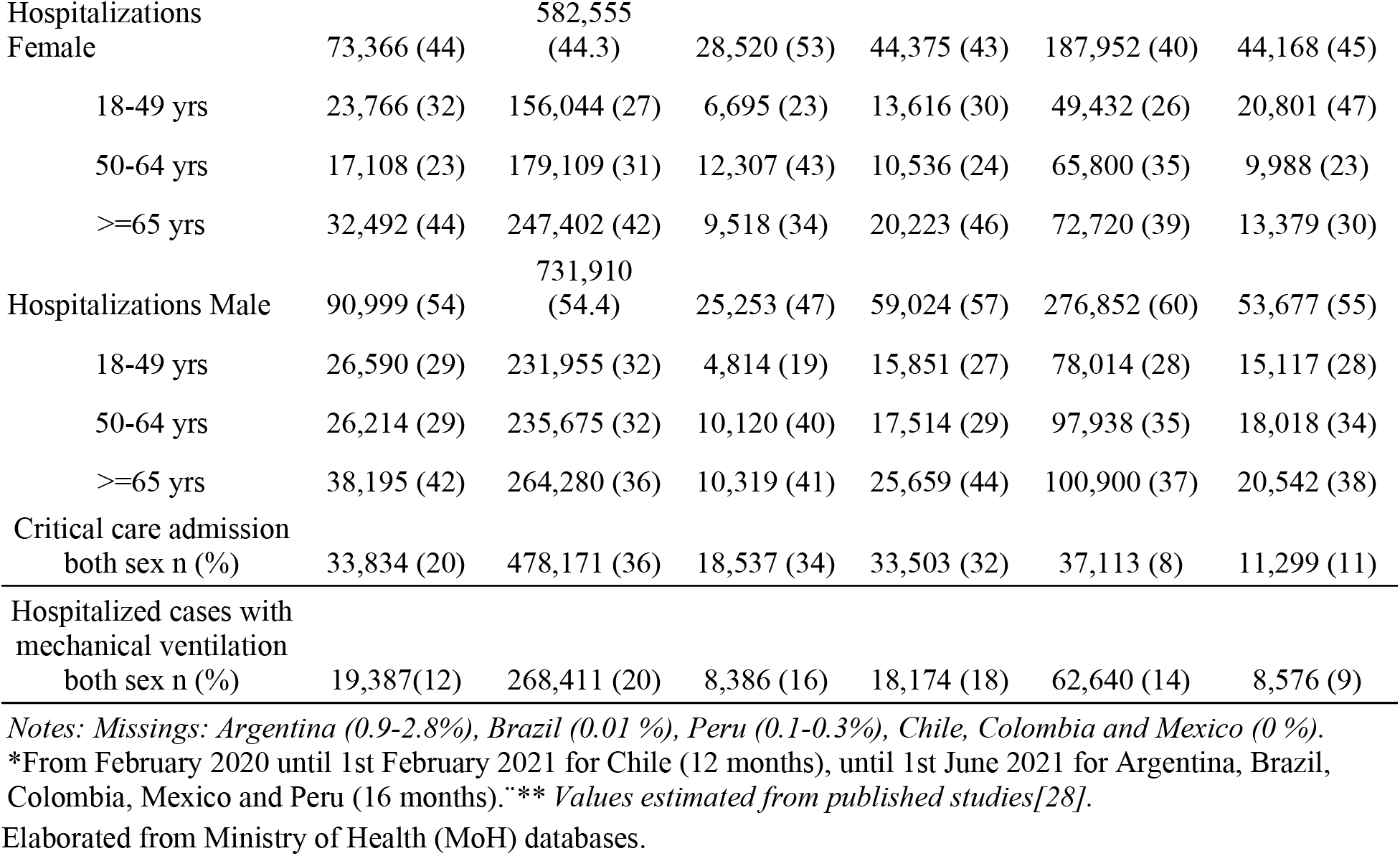
COVID-19 Hospitalization admission by country per sex and age, critical care admission, and hospitalized cases with mechanical ventilation*.

Data on COVID-19 individuals with certain comorbidities are summarized in Table S20 of the Supplementary Material. The majority of the analyzed cohorts had diabetes (7.5 to 23.2%) or hypertension (14.4 to 40.8%) as associated comorbidities in all countries. Brazil reported the largest proportion of COVID-19 cases with diabetes (20.3%), whereas Colombia reported the largest proportion of cases with hypertension (40.8%).

### Mortality

A total of 1,144,544 people over 18 years of age died of COVID-19 during the selected period, representing 5% of the overall cases. Even though the majority of deaths were male (59%), when stratified by age and sex, a larger proportion of females over 65 years of age died than males of the same age group. Table 3 summarizes death counts and percentages by country, sex, and age. The case fatality rate during the pre-vaccination period varied between countries from 2.5% (Argentina and Chile) to 10% (Mexico and Peru) (Table 3). As shown in Figure S8 in the Supplemental Material, the monthly CFR trajectories revealed elevated CFR rates for Mexico and Peru, confirming the global CFR data.

**Table 3.** COVID-19 Death counts and percentages by country, sex and age*.

| Indicators n (%) | Argentina | Brazil | Chile | Colombia | Mexico | Peru |
| --- | --- | --- | --- | --- | --- | --- |
| Death both sex | 92,434 (100) | 521,577 (100) | 18,480 (100) | 89,137 (100) | 237,947 (100) | 184,969 (100) |
| 18-49 yrs | 6,816 (7) | 88,035 (16.9) | 1,041 (5.6) | 8,928 (10) | 37,678 (16) | 23,947 (13) |
| 50-64 yrs | 19,257 (21) | 140,505<br>(26.9) | 5,679 (30.7) | 22,104 (25) | 82,553 (35) | 54,055 (29) |
| >=65 yrs | 66,361 (72) | 293,037<br>(56.2) | 11,760 (63.6) | 58,105 (65) | 117,716 (49) | 106,967 (58) |
| Death Female | 37,714 (41) | 228,360<br>(43.8) | 6,542 (35.4) | 33,978 (38) | 89,116 (37) | 66,805 (36) |
| 18-49 yrs | 2,531 (7) | 35,620 (15.6) | 262 (4) | 2,912 (9) | 12,074 (14) | 7,729 (12) |
| 50-64 yrs | 6,507 (17) | 58,070 (25.4) | 2,682 (40.9) | 7,834 (23) | 30,249 (34) | 18,360 (27) |
| >=65 yrs | 28,676 (76) | 134,670<br>(58.9) | 3,598 (54.9) | 23,232 (68) | 46,793 (52) | 40,716 (61) |
| Death Male | 52,728 (57) | 293,159 (50) | 11,938 (64.6) | 55,159 (62) | 148,831 (63) | 118,164 (64) |
| 18-49 yrs | 4,201 (8) | 52,402 (18) | 477(4) | 6,016 (11) | 25,604 (17) | 16,218 (14) |
| 50-64 yrs | 12,604 (24) | 82,419 (28) | 4,895 (41) | 14,270 (26) | 52,304 (35) | 35,695 (30) |
| >=65 yrs | 35,923 (68) | 158,338 (54) | 6,566 (55) | 34,873 (63) | 70,923 (48) | 66,251 (56) |
| Mortality rate<br>per 100000 | 276.1 | 327.5 | 123.4 | 238.8 | 271.8 | 814.9 |
| Case fatality rate | 2.5 | 4.4 | 2.5 | 2.8 | 10.1 | 9.9 |

Notes: Missing: Argentina (0.9- 2.7%), Brazil (both sex: 0.01-0.02%), Chile, Colombia, Mexico and Peru (0%). \*from February 2020 until 1st February 2021 for Chile (12 months), until 1st June 2021 for Argentina, Brazil, Colombia, Mexico and Peru (16 months)). Elaborated from Ministry of Health (MoH) databases.

### Years of life lost and excess mortality

We identified and prioritized five studies that estimated the burden of COVID-19 and used the DALYs and/or YLLs as metrics (Table S2). The overall analyses of YLLs primarily rely on reports from multiple countries based on ministerial databases, encompassing different analysis periods throughout 2020, ranging from eight months to one year (Table S3). There was high variability in the estimation of YLL, ranging from 241,089 in Chile to 3,312,346 in Brazil (Table 6). Peru and Mexico had the highest DALYs per 100,000 people, ranging from 2,549.5 and 3,510.7.

**Table 6.** Years of life lost (YLLs) and disability-adjusted life years (DALYs) per 100,000 persons in six countries*.

| Country | YLLs | YLDs | DALYs | DALYs/100,000 |
| --- | --- | --- | --- | --- |
| Argentina | 510,222 | 9,235 | 519,457 | 1,680.6 |
| (min-max) | - | (3,418 - 69,900) | (513,640 - 580,122) | (1,661 - 1,876) |
| Brazil | 3,312,346 | 59,953 | 3,372,299 | 2,209 |
| (min-max) | - | (22,192 - 453,791) | (3,334,538 - 3,766,137) | (2,184 - 2,467) |
| Chile | 241,089 | 4,363 | 245,452 | 1,697.3 |
| (min-max) | - | (1,615 - 33,029) | (242,704 - 274,118) | (1,678 - 1,895) |
| Colombia | 885,793 | 16,033 | 901,826 | 2,532.7 |
| (min-max) | - | (5,934 - 121,353) | (891,727 - 1,007,146) | (2,504 - 2,828) |
| Mexico | 2,097,504 | 37,761 | 2,135,265 | 2,549.5 |
| (min-max) | - | - | - | - |
| Peru | 744,331 | 13,472 | 757,803 | 3,510.7 |
| (min-max) | - | (4,987 - 101,973) | (749,318 - 846,304) | (3,471 - 3,920) |

Notes: YLL data were retrieved from the studies shown in Table S2. All Years Lived with Disability (YLD) estimates were derived using the ratio (YLL/YLD) calculated in the study conducted by Escudero-Salinas and multiplied by the YLL of each country.[29] The Min-Max values for estimating Disability-Adjusted Life Years (DALYs) were determined by utilizing the range of values reported in a systematic review [30; the] YLDs range was calculated from the DALYs range and YLL.\*Period under pre-vaccination analysis. Elaborated from Ministry of Health (MoH) databases.

Based on the data provided in these reports, we estimated age-standardized YLL for the six Latin American countries stratified by age group (Figure 3). For more details, see Tables S4-9 in the Supplemental Material. The age-standardized YLL per 100,000 persons ranged from 945 in Argentina to 2,268 in Peru. Younger patients (<65 years) contributed more to YLLs rates than adults aged > 65 years.

**Figure 3.**
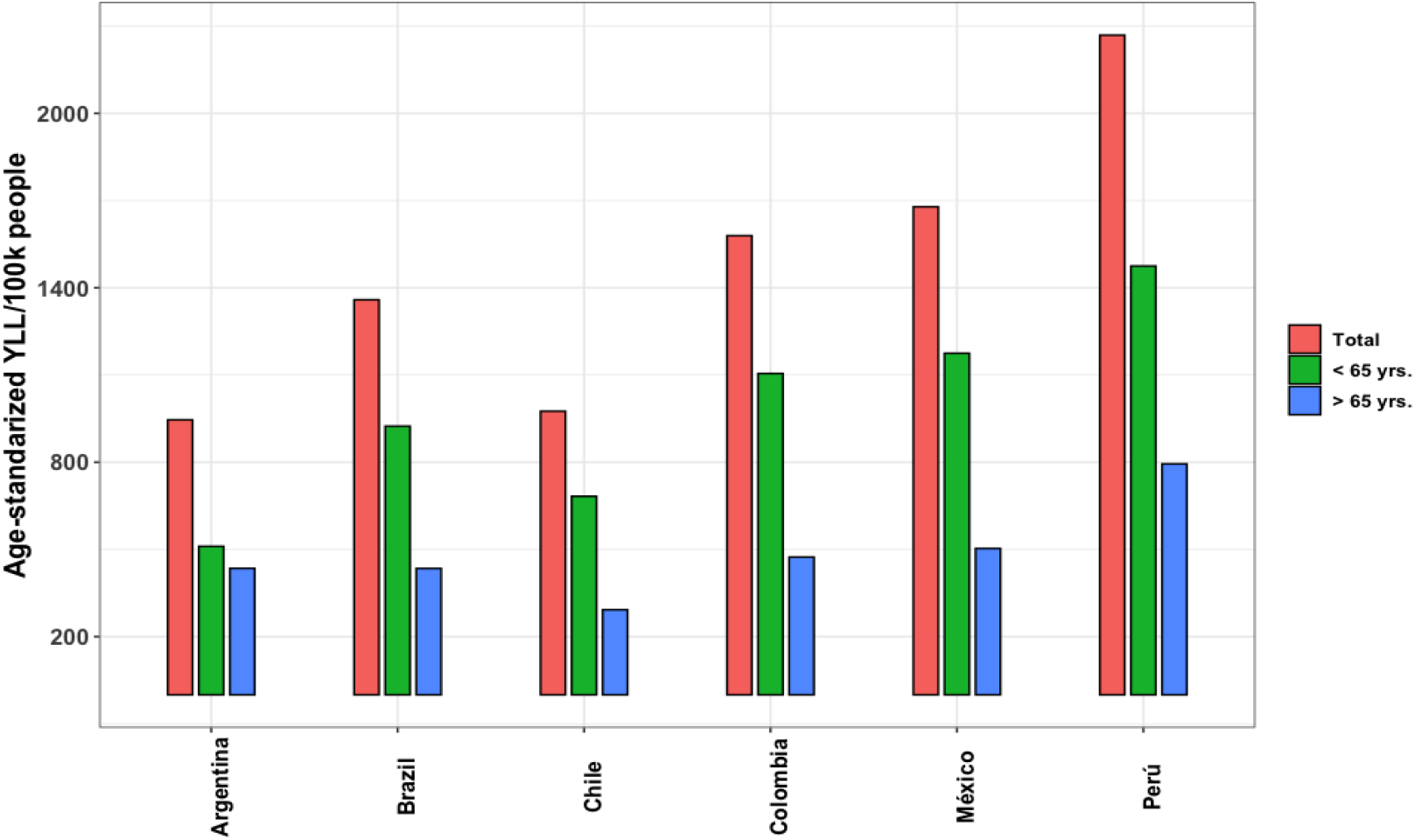
Total age-standardized years of life lost rates for COVID-19 by country and age group. **\*** Notes: using data from Salinas-Escudero, 98.6% of total YLL rates were employed to estimate age-standardized YLL rates for individuals aged 20 years and above.[29]. Source: All data were retrieved from the studies shown in Table S2. *Period under analysis pre-vaccination

Regarding excess mortality, we prioritized the five papers identified through a systematic review. The characteristics of the included studies are shown in Table S2. The analysis period encompassed almost all cases in 2020; however, significant variations in excess mortality were observed, ranging from a p-score of 10.6 to 109.5 (Table S10). The total excess mortality per 100,000 people ranged from 76 to 557 in Argentina and Peru, respectively (Figure 4). In Argentina, Brazil, and Chile, COVID-19 deaths have exceeded excess mortality, while in Colombia, Mexico, and Peru, excess mortality rates have surpassed those of COVID-19 deaths. Regarding age, excess mortality data were accessible only for Mexico and Brazil (Supplementary Table S11), revealing that the overall excess mortality and the proportion of excess deaths attributable to COVID-19 were greater among males than females.

**Figure 4.**
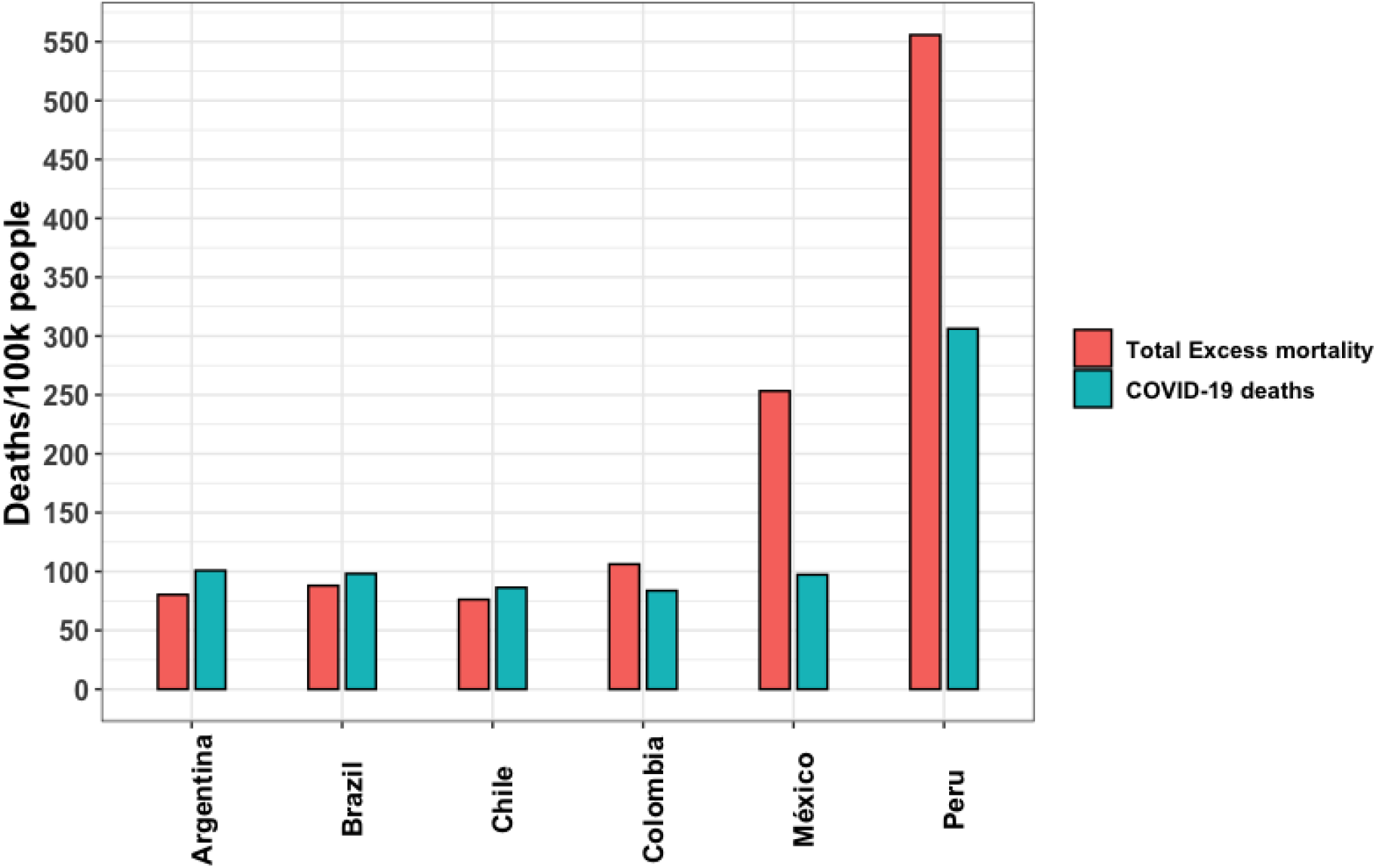
All age ranges of total excess mortality and COVID-19 deaths per 100,000 persons in six Latin American countries in 2020. Source: Author’s elaboration based on the literature review.

### Economic burden of COVID-19

Table 8 details the direct medical costs related to COVID-19, classifying them according to the cost components and disease severity in specific countries. The costs per case of mild COVID-19 ranged from USD 26.6 in Peru to USD 68.9 in Argentina. In mild cases, diagnosis and laboratory tests represent the resource with the greatest share of the total cost in Argentina, Brazil, Mexico, and Peru. By contrast, in Chile and Colombia, medical consultations are the largest component of the cost per mild case.

**Table 8.** Cost per case of COVID 19 in US Dollars*.

|  | Argentina | Brazil | Chile | Colombia | Mexico | Peru |
| --- | --- | --- | --- | --- | --- | --- |
| Mild | \$68.9 (100.0%) | \$26.6 (100.0%) | \$56.3 (100.0%) | \$28.9 (100.0%) | \$44.2 (100.0%) | \$27.6 (100.0%) |
| Consultations | \$20.0 (29.0%) | \$7.2 (27.1%) | \$46.5 (82.5%) | \$13.9 (48.2%) | \$20.5 (46.5%) | \$13.2 (47.7%) |
| Diagnostic and laboratory tests | \$45.5 (66.0%) | \$17.6 (66.1%) | \$9.6 (17.0%) | \$13.1 (45.3%) | \$21.9 (49.6%) | \$14.1 (50.9%) |
| Hospitalizations | \$0.0 (0.0%) | \$0.0 (0.0%) | \$0.0 (0.0%) | \$0.0 (0.0%) | \$0.0 (0.0%) | \$0.0 (0.0%) |
| Drugs | \$3.4 (5.0%) | \$1.8 (6.8%) | \$0.3 (0.6%) | \$1.9 (6.5%) | \$1.8 (4.0%) | \$0.4 (1.4%) |
| Moderate and severe | \$2,510.0 (100.0%) | \$1,059.4 (100.0%) | \$2,971.3 (100.0%) | \$1,721.8 (100.0%) | \$1,936.6 (100.0%) | \$1,357.7 (100.0%) |
| Consultations | \$9.1 (0.4%) | \$2.8 (0.3%) | \$24.3 (0.8%) | \$12.2 (0.7%) | \$12.2 (0.6%) | \$7.8 (0.6%) |
| Diagnostic and laboratory tests | \$242.3 (9.7%) | \$77.0 (7.3%) | \$279.4 (9.4%) | \$448.3 (26.0%) | \$311.0 (16.1%) | \$199.8 (14.7%) |
| Hospitalizations | \$2,253.0 (89.8%) | \$976.6 (92.2%) | \$2,667.1 (89.8%) | \$1,258.2 (73.1%) | \$1,610.0 (83.1%) | \$1,149.5 (84.7%) |
| Drugs | \$5.7 (0.2%) | \$3.0 (0.3%) | \$0.5 (0.0%) | \$3.1 (0.2%) | \$3.5 (0.2%) | \$0.6 (0.0%) |
| Critical | \$23,384.2 (100.0%) | \$19,391.5 (100.0%) | \$19,839.8 (100.0%) | \$7,147.2 (100.0%) | \$30,040.9 (100.0%) | \$8,053.8 (100.0%) |
| Consultations | \$9.2 (0.0%) | \$2.8 (0.0%) | \$25.2 (0.1%) | \$12.7 (0.2%) | \$12.6 (0.0%) | \$8.1 (0.1%) |
| Diagnostic and laboratory tests | \$1,207.7 (5.2%) | \$351.4 (1.8%) | \$1,654.2 (8.3%) | \$2,280.8 (31.9%) | \$1,606.5 (5.3%) | \$1,032.1 (12.8%) |
| Hospitalizations | \$8,611.8 (36.8%) | \$5,217.6 (26.9%) | \$9,211.8 (46.4%) | \$3,806.7 (53.3%) | \$5,936.1 (19.8%) | \$4,911.5 (61.0%) |
| Drugs | \$13,555.5 (58.0%) | \$13,819.7 (71.3%) | \$8,948.7 (45.1%) | \$1,047.1 (14.7%) | \$22,485.8 (74.9%) | \$2,102.1 (26.1%) |

Source: own elaboration
\*Cost reported for the year 2023

The costs for moderate and severe cases vary between USD 1,059.4 in Brazil and USD 2,951.3 in Chile, with hospitalization being the resource with the greatest weight in the cost for this type of case in all the countries analyzed.

On the other hand, the costs per critical case ranged between USD 7,147.2 in Colombia and USD 30,040.9 in Mexico. In Argentina, Brazil, and Mexico, the resource with the greatest impact in these cases is related to drug spending, whereas in Chile, Colombia, and Peru, the most significant component is associated with hospitalization.

Therefore, the total cost associated with COVID-19 ranged from $250 million in Peru to $10 billion in Brazil. This represents 2% of the total health spending per country, except for Brazil, where it represents 5% (See Table 9). Critical cases represent the largest proportion of the total costs due to COVID-19. However, in the case of Peru, moderate and severe cases contribute more to the total cost, given that the proportion of critical cases in Peru (0.6% of total COVID cases) is relatively lower than in other countries.

**Table 9.**
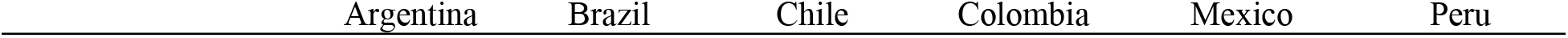
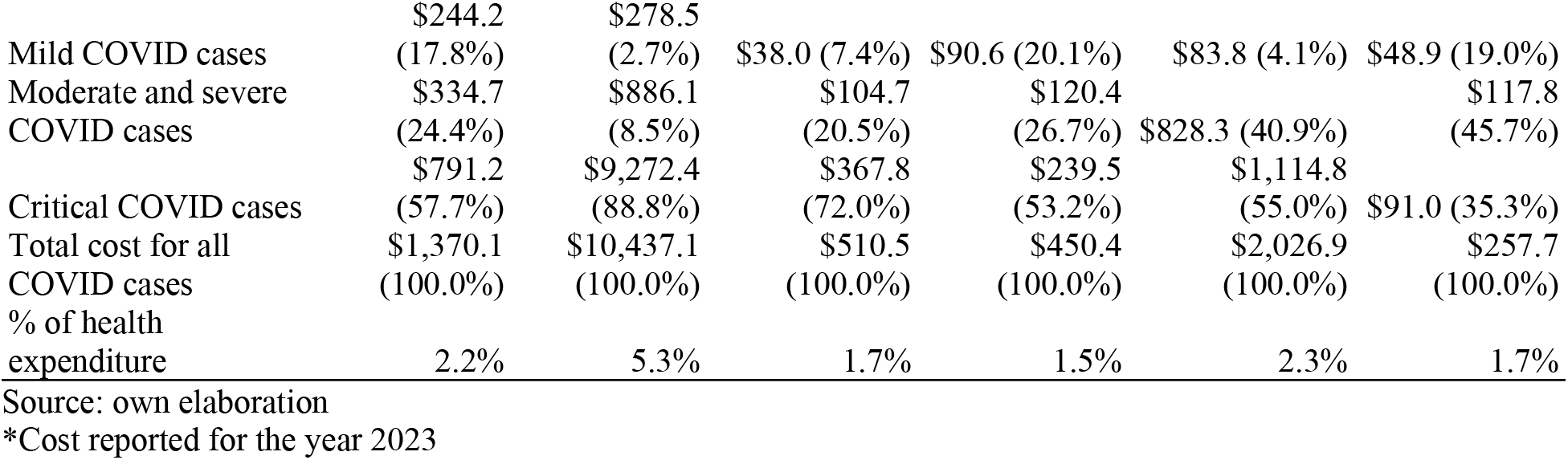
Total cost of COVID-19 by severity in US Dollars*.

### Other vaccine-preventable diseases

Data from pre-vaccinal cases of pneumonia and influenza in the selected countries presented substantial heterogeneity, probably given the different onset of vaccination programs and dated of the data. Table S16 in Supplementary Material summarized the average death counts of pneumonia and influenza. Argentina, along with Brazil, experienced the highest number of deaths compared to the other countries.

The costs per case of hospitalization due to pneumococcus found in studies vary between $800 and $1900 in Argentina [31], Brazil [32], and Colombia [33–35]. On the other hand, three studies from Argentina [36,37] and Peru [38] were identified for the costs per case of hospitalization due to influenza that reported costs between 20$ and $400. Further details on the results regarding other vaccine-preventable diseases are presented in Supplementary Material 8.

## Discussion

In this study, we provided a comprehensive overview of the COVID-19 burden in six countries of the Latin America region before implementation of the nationwide vaccination program. These countries lead the economic income of the region and represent approximately 80% of the Latin American population. Our main finding was a substantial disease burden across all six countries, with variations in all the outcomes considered.

We observed a high degree of variability in disease burden (incidence, hospitalization, death, excess mortality, and years of life lost), as well as direct medical costs could be attributed to the diverse strategies employed by each country during the peak of the pandemic. These strategies include factors such as mandatory testing in hospital settings, delays in test results, discrepancies in case reporting, and variations in case definitions, particularly before the availability of vaccines.[39] In addition, the availability of healthcare resources and treatment is also likely to impact these estimates. Crossing the six countries, most showed mild disease rates close to 90%, whereas severe disease rates were less than 4%. It is crucial to note that these proportions can be influenced by surveillance methods, therapeutic use, interventions, regional demographics, vaccination rates, and emerging variants. In national databases, assigning severity categories and determining hospital or ICU admissions could depend on various factors, including clinical judgment and local and national policies, rather than prognosis prediction models.[40]

Another remarkable result was that in most cases, the percentage of patients needing mechanical ventilation was nearly half of those requiring critical care. However, this relationship was more pronounced in Peru and Mexico, with 80% for Peru and almost twice as high for Mexico. One plausible explanation is that hospitals had to provide mechanical ventilation outside traditional ICUs due to high demand. This was evident early in the pandemic, before widespread vaccination. In a Mexican cohort study, 45% of non-survivors with critical illness requiring ICU admission did not receive Invasive Mechanical Ventilation (IMV) or ICU care due to limited ICU beds. A similar scenario was reported for the entire Metropolitan area of Mexico City, indicating a shortage of ICU beds and hospital overcrowding, leading to delayed ICU admissions.[41]

Regarding mortality indicators, Peru experienced the most severe situation, a fact corroborated by other authors.[42] This situation likely stemmed from the health system having limited capacity and fewer resources to cope with the unfolding pandemic, as reported in other regions as well.[43,44] However, it should be noted that the Peruvian government, at the end of the first wave, changed the method of recording deaths, almost doubling the registered deaths with the new, more sensitive method, which may have influenced these significant differences.[45]

The similarity in values between YLLs and DALYs was primarily due to the fact that the main component of DALYs was mortality, while the disability weight associated with the acute phase of the disease was lower.[46] On the other hand, in countries where COVID-19 deaths exceeded excess mortality, one possible reason may be the decrease in mortality from different causes (such as trauma, seasonal viruses and cardiovascular problems) due to the mobility restrictions imposed and the isolation measures adopted by policy makers;[47] while in countries where the opposite is the case, one possible explanation could be the tensions in the health system that limit access to conditions such as cancer and heart disease.[47,48] Underreporting, especially in LMICs, may widen this gap.[45]

In economic terms, this study reveals that before vaccination implementation, total direct medical costs related to COVID-19 represented approximately 2% of total health spending in most cases, except Brazil (5%). A study on the economic burden of COVID-19 in Iran was found [49], which concluded that the total cost in 2020 was equivalent to approximately 7% of health spending. On the other hand, an analysis carried out in Saudi Arabia between March 2020 and January 2021, in a sample of 5,286 patients [50], revealed that the medical costs of ICU hospitalization per patient averaged USD 4,580. Comparatively, in our study, critically ill patients requiring ICU care had a total hospitalization cost of between USD 3,807 and USD 8,612.

This study displays several strengths. Firstly, the thoroughness of the search process is a notable asset. We meticulously scoured official sources to collect pertinent data regarding the impact of COVID-19 on six countries representing almost 80% of the Latin American population. Secondly, the use of systematic review to search for the best evidence as a source for calculating EM and YLL coupled with the supplementation of official databases, represents another notable strength. These two outcomes offer a more comprehensive and conclusive understanding of mortality aspects, especially given the inherent challenges in analyzing mortality data in the context of an outbreak/pandemic. Finally, the cost analysis addresses a crucial aspect of the disease burden, particularly in low-middle-income countries.

This study has certain limitations. First, a limitation arises from the lack of information and heterogeneity of data recording in the countries. Nevertheless, the research team used different methods to homogenize this information. However, it is important to note that, despite these adjustments, a comparison between countries is not ideal. Second, we assessed the disease burden during the dominance of Alpha and Beta variants in a predominantly unvaccinated population without addressing the spread of variants with varying degrees of severity within the region. On the other hand, the cost estimation is based on an approximation based on WHO clinical guidelines adapted by local experts, which may have the limitation of not accurately representing the reality of the countries. In addition, the cost information was extracted from different sources, which may bias the comparison between countries. However, this methodology is commonly used because of the lack of a health system resource use registry and cost of illness studies in the region. This shows the need for more studies examining the impact of COVID-19 on the use of healthcare resources and related costs in the adult population. Finally, YLL (and related measures) must be used carefully regarding COVID-19, a disease that primarily affects the elderly. More extensive longevity in a population results in a higher YLL, as observed in Cuba. This effect persists even with age-adjusted YLL. For illustration, consider an extreme case where half of the people over 70 years of age die from a disease. In a population with no individuals over 70 years of age, the age-adjusted YLL was 0. In contrast, the YLL was positive in the other with a significantly older population. This effect cannot be fully mitigated with age-adjusted YLL. Nevertheless, the figures presented represent a helpful starting point.

## Conclusion

The COVID-19-related health and economic burden was significant and exhibited considerable variability among the six Latin American countries during the pre-vaccination period. This study highlights several key areas that require attention in the Latin American region. First, there is a need to standardize and improve the definitions used in health information systems. The lack of uniformity in definitions can lead to inconsistencies in data interpretation, making cross-country comparisons difficult and affecting evidence-based decision making. Second, it is crucial to standardize data collection and management practices. The heterogeneity in methods used by different regional countries and entities presents significant challenges for regional data integration and analysis. Implementing data collection and management standards can facilitate data comparison and cross-national collaboration.

## Data Availability

All data involved in this study are included in the main manuscript and its supplementary information files.

## Ethical Conduct of the Study

This is a review of available published data. Therefore, no ethical considerations were needed.

## Author Contributions

All authors contributed to the development of the publication and maintained control over the final content.

## References

1. Schwalb A, Armyra E, Méndez-Aranda M, Ugarte-Gil C. COVID-19 in Latin America and the Caribbean: Two years of the pandemic. J Intern Med. 2022;292:409–427.

2. Vargas ED, Sanchez GR. COVID-19 Is Having a Devastating Impact on the Economic Well-being of Latino Families. Journal of Economics, Race, and Policy. 2020;3:262–269.

3. Diseases C. Manejo clínico de la COVID-19: orientaciones evolutivas, 25 de enero de 2021. World Health Organization; 23 de noviembre de 2021 [cited 13 Mar 2024]. Available: https://www.who.int/es/publications/i/item/WHO-2019-nCoV-clinical-2021-1

4. Martinez R, Soliz P, Caixeta R, Ordunez P. Reflection on modern methods: years of life lost due to premature mortality-a versatile and comprehensive measure for monitoring non-communicable disease mortality. Int J Epidemiol. 2019;48. doi:10.1093/ije/dyy254

5. Msemburi W, Karlinsky A, Knutson V, Aleshin-Guendel S, Chatterji S, Wakefield J. The WHO estimates of excess mortality associated with the COVID-19 pandemic. Nature. 2022;613:130–137.

6. Mogyorosy Z, Smith PC. The Main Methodological Issues in Costing Health Care Services - a Literature Review. 2005. Available: https://pure.york.ac.uk/portal/en/publications/the-main-methodological-issues-in-costing-health-care-services-a--2.

7. WHO. Living guidance for clinical management of COVID-19. In: World Health Organization [Internet]. Available: https://apps.who.int/iris/bitstream/handle/10665/349321/WHO-2019-nCoV-clinical-2021.2-eng.pdf

8. Johnston RJ, Rolfe J, Rosenberger RS, Brouwer R. Introduction to Benefit Transfer Methods. Benefit Transfer of Environmental and Resource Values. 2015; 19–59.

9. Rosenberger RS, Stanley T. Measurement, generalization, and publication: Sources of error in benefit transfers and their management. Ecol Econ. 2006;60:372–378.

10. Navrud S, Ready R. Review of Methods for Value Transfer. Environmental Value Transfer: Issues and Methods. 2007; 1–10.

11. SIGTAP - Sistema de Gerenciamento da Tabela de Procedimentos, Medicamentos e OPM do SUS. [cited 25 Oct 2023]. Available: http://sigtap.datasus.gov.br/tabela-unificada/app/sec/inicio.jsp?first=15

12. Consultorsalud. Manual Tarifario de Salud SOAT 2023 - versión PDF. In: CONSULTORSALUD [Internet]. 2 Jan 2023 [cited 26 Oct 2023]. Available: https://consultorsalud.com/manual-tarifario-de-salud-soat-2023-version-pdf/

13. Website. Available: MinSalud. ACUERDO No. 256 DE 2001. Ministerio de Salud y Protección Social https://lexsaludcolombia.files.wordpress.com/2010/10/tarifas-iss-2001.pdf.

14. Collao CG. Eventual Reforma de FONASA Hacia Una Particular ISAPR. Pública, Con Dos Categorías Más FONASA PLUS.”. Observatorio Chileno de Salud Pública; 2023. Available: https://www.ochisap.cl/wp-content/uploads/2023/04/Reforma-de-FONASA-2023-C.-Gattini-OCHISAP.pdf

15. kairos - El portal farmacéutico. [cited 26 Oct 2023]. Available: https://kairosweb.com/

16. Minsalud. Termómetro de precios de medicamentos. [cited 26 Oct 2023]. Available: https://www.minsalud.gov.co/salud/MT/Paginas/termometro-de-precios.aspx

17. Minsa. Observatorio Peruano de Productos Farmacéuticos. In: SISTEMA NACIONAL DE INFORMACIÓN DE PRECIOS DE PRODUCTOS FARMACÉUTICOS - SNIPPF [Internet]. [cited 13 Oct 2023]. Available: https://opm-digemid.minsa.gob.pe/#/consulta-producto

18. Extended National Consumer Price Index. [cited 25 Oct 2023]. Available: https://www.ibge.gov.br/en/statistics/economic/prices-and-costs/17129-extended-national-consumer-price-index.html?=&t=series-historicas

19. INDEC, Instituto Nacional de Estadistica y Censos de la REPUBLICA ARGENTINA. INDEC: Instituto Nacional de Estadística y Censos de la República Argentina. [cited 30 Oct 2023]. Available: https://www.indec.gob.ar/

20. DANE - IPC información técnica. [cited 25 Oct 2023]. Available: https://www.dane.gov.co/index.php/estadisticas-por-tema/precios-y-costos/indice-de-precios-al-consumidor-ipc/ipc-informacion-tecnica

21. Chile INE. Calculadora IPC. In: Instituto Nacional de Estadística -INE-, Chile [Internet]. [cited 26 Oct 2023]. Available: https://calculadoraipc.ine.cl/

22. Banco Central de Reserva del Perú. [cited 30 Oct 2023]. Available: https://www.bcrp.gob.pe/

23. Banco Central de la Repblica Argentina. [cited 13 Mar 2024]. Available: https://www.bcra.gob.ar/

24. Banco Central do Brasil. [cited 13 Mar 2024]. Available: https://www.bcb.gov.br/conversao

25. Banco Central de Chile. In: Banco Central de Chile [Internet]. [cited 13 Mar 2024]. Available: https://www.bcentral.cl/inicio

26. Banco de la República de Colombia. [cited 13 Mar 2024]. Available: https://www.banrep.gov.co/es

27. Mathieu E, Ritchie H, Rodés-Guirao L, Appel C, Giattino C, Hasell J, et al. Coronavirus Pandemic (COVID-19). Our World in Data. 2020 [cited 4 Oct 2023]. Available: https://ourworldindata.org/coronavirus

28. Araujo M, Ossandón P, Abarca AM, Menjiba AM, Muñoz AM. [Prognosis of patients with COVID-19 admitted to a tertiary center in Chile: A cohort study]. Medwave. 2020;20:e8066.

29. Salinas-Escudero G, Toledano-Toledano F, García-Peña C, Parra-Rodríguez L, Granados-García V, Carrillo-Vega MF. Disability-adjusted life years for the COVID-19 pandemic in the Mexican population. Front Public Health. 2021;9:686700.

30. Gebeyehu DT, East L, Wark S, Islam MS. Disability-adjusted life years (DALYs) based COVID-19 health impact assessment: a systematic review. BMC Public Health. 2023;23:1–13.

31. Giglio ND, Castellano VE, Mizrahi P, Micone PV. Cost-Effectiveness of Pneumococcal Vaccines for Adults Aged 65 Years and Older in Argentina. Value in health regional issues. 2022;28. doi:10.1016/j.vhri.2021.08.003

32. Michelin L, Weber FM, Scolari BW, Menezes BK, Gullo MC. Mortality and costs of pneumococcal pneumonia in adults: a cross-sectional study. J Bras Pneumol. 2019;45. doi:10.1590/1806-3713/e20180374

33. Castañeda-Orjuela C, Alvis-Guzmán N, Paternina AJ, la Hoz-Restrepo FD. Cost-effectiveness of the introduction of the pneumococcal polysaccharide vaccine in elderly Colombian population. Vaccine. 2011;29. doi:10.1016/j.vaccine.2011.08.006

34. Pugh S, Wasserman M, Moffatt M, Marques S, Reyes JM, Prieto VA, et al. Estimating the Impact of Switching from a Lower to Higher Valent Pneumococcal Conjugate Vaccine in Colombia, Finland, and The Netherlands: A Cost-Effectiveness Analysis. Infectious Diseases and Therapy. 2020;9:305– 324.

35. Ordóñez JE, Ordóñez A. A cost-effectiveness analysis of pneumococcal conjugate vaccines in infants and herd protection in older adults in Colombia. Expert Rev Vaccines. 2023;22. doi:10.1080/14760584.2023.2184090

36. Urueña A, Micone P, Magneres C, Mould-Quevedo J, Giglio N. Cost-Effectiveness Analysis of Switching from Trivalent to Quadrivalent Seasonal Influenza Vaccine in Argentina. Vaccines. 2021;9:335.

37. Cost-effectiveness of introducing an MF59-adjuvanted trivalent influenza vaccine for older adults in Argentina. Vaccine. 2020;38:3682–3689.

38. Tinoco YO, Azziz-Baumgartner E, Rázuri H, Kasper MR, Romero C, Ortiz E, et al. A population-based estimate of the economic burden of influenza in Peru, 2009–2010. Influenza Other Respi Viruses. 2016;10:301–309.

39. Burki T. COVID-19 in Latin America. Lancet Infect Dis. 2020;20:547–548.

40. Diseases C. Living guidance for clinical management of COVID-19. World Health Organization; 23 Nov 2021 [cited 13 Mar 2024]. Available: https://www.who.int/publications/i/item/WHO-2019-nCoV-clinical-2021-2

41. Olivas-Martínez A, Cárdenas-Fragoso JL, Jiménez JV, Lozano-Cruz OA, Ortiz-Brizuela E, Tovar-Méndez VH, et al. In-hospital mortality from severe COVID-19 in a tertiary care center in Mexico City; causes of death, risk factors and the impact of hospital saturation. PLoS One. 2021;16:e0245772.

42. Ramírez-Soto MC, Ortega-Cáceres G. Analysis of Excess All-Cause Mortality and COVID-19 Mortality in Peru: Observational Study. Tropical Medicine and Infectious Disease. 2022;7:44.

43. Merlo F, Lepori M, Malacrida R, Albanese E, Fadda M. Physicians’ acceptance of the Swiss Academy of Medical Sciences guidelines “COVID-19 pandemic: triage for intensive-care treatment under resource scarcity.” Swiss Med Wkly. 2021;151:w20472.

44. Liu X, Li X, Sun T, Qin H, Zhou Y, Zou C, et al. East-West differences in clinical manifestations of COVID-19 patients: A systematic literature review and meta-analysis. J Med Virol. 2021;93:2683– 2693.

45. Valdez Huarcaya W, Miranda Monzón JA, Napanga Saldaña EO, Driver CR. Impacto de la COVID-19 en la mortalidad en Perú mediante la triangulación de múltiples fuentes de datos. en la mortalidad en Perú mediante la triangulación de múltiples fuentes de datos Rev Panam Salud Publica. 2022;2022:e53.

46. Gianino MM, Savatteri A, Politano G, Nurchis MC, Pascucci D, Damiani G. Burden of COVID-19: Disability-Adjusted Life Years (DALYs) across 16 European countries. Eur Rev Med Pharmacol Sci. 2021;25:5529–5541.

47. dos Santos AM, de Souza BF, de Carvalho CA, Campos MAG, de Oliveira BLCA, Diniz EM, et al. Excess deaths from all causes and by COVID-19 in Brazil in 2020. Rev saúde pública. 2021;55:71– 71.

48. Mejía LSP, Fernández JLW, Hernández IO, Ridaura RL, Ramirez HL-G, Avila MH, et al. Estimación del exceso de mortalidad por todas las causas durante la pandemia del Covid-19 en México. Salud Publica Mex. 2021;63:211–224.

49. Ghaffari Darab M, Keshavarz K, Sadeghi E, Shahmohamadi J, Kavosi Z. The economic burden of coronavirus disease 2019 (COVID-19): evidence from Iran. BMC Health Serv Res. 2021;21:1–7.

50. Alenzi KA, Al-malky HS, Altebainawi AF, Abushomi HQ, Alatawi FO, Atwadi MH, et al. Health economic burden of COVID-19 in Saudi Arabia. Front Public Health. 2022;10:927494.

